# Development of a predictive risk model for severe COVID-19 disease using population-based administrative data

**DOI:** 10.1101/2020.10.21.20217380

**Authors:** Jiandong Zhou, Sharen Lee, Xiansong Wang, Yi Li, William KK Wu, Tong Liu, Zhidong Cao, Daniel Dajun Zeng, Ian Chi Kei Wong, Bernard Man Yung Cheung, Qingpeng Zhang, Gary Tse

**Author notes:** Prof. Gary Tse, PhD, FRCP, FFPH*, Tianjin Key Laboratory of Ionic-Molecular Function of Cardiovascular Disease, Department of Cardiology, Tianjin Institute of Cardiology, Second Hospital of Tianjin Medical University, Tianjin 300211, China, *Prof. Qingpeng Zhang, PhD*, School of Data Science, City University of Hong Kong, Hong Kong, China.

## Abstract

**Background:** Recent studies have reported numerous significant predictors for adverse outcomes in COVID-19 disease. However, there have been few simple clinical risk score for prompt risk stratification. The objective is to develop a simple risk score for severe COVID-19 disease using territory-wide healthcare data based on simple clinical and laboratory variables.

**Methods:** Consecutive patients admitted to Hong Kong’s public hospitals between 1^st^ January and 22^nd^ August 2020 diagnosed with COVID-19, as confirmed by RT-PCR, were included. The primary outcome was composite intensive care unit admission, need for intubation or death with follow-up until 8^th^ September 2020.

**Results:** COVID-19 testing was performed in 237493 patients and 4445 patients (median age 44.8 years old, 95% CI: [28.9, 60.8]); 50% male) were tested positive. Of these, 212 patients (4.8%) met the primary outcome. A risk score including the following components was derived from Cox regression: gender, age, hypertension, stroke, diabetes mellitus, ischemic heart disease/heart failure, respiratory disease, renal disease, increases in neutrophil count, monocyte count, sodium, potassium, urea, alanine transaminase, alkaline phosphatase, high sensitive troponin-I, prothrombin time, activated partial thromboplastin time, D-dimer and C-reactive protein, as well as decreases in lymphocyte count, base excess and bicarbonate levels. The model based on test results taken on the day of admission demonstrated an excellent predictive value. Incorporation of test results on successive time points did not further improve risk prediction.

**Conclusions:** A simple clinical score accurately predicted severe COVID-19 disease, even without including symptoms, blood pressure or oxygen status on presentation, or chest radiograph results.

## Introduction

The coronavirus disease 2019 has a wide clinical spectrum, with disease severities ranging from completely asymptomatic to the need for intubation and death. For example, those with existing cardiac problems are more likely to suffer from more severe disease life courses (Guo et al., 2020;Li et al., 2020;Shi et al., 2020;Wang et al., 2020). Aside from comorbidities, numerous risk factors such as high D-dimer (Yao et al., 2020), neutrophil (Huang et al., 2020), and liver damage (Cai et al., 2020) and deranged clotting (Tang et al., 2020) have been associated with disease severity. Such patients may benefit from early aggressive treatment. However, to date there are only a few easy-for-use risk models that can be used for early identification of such at-risk individuals in clinical practice (Knight et al., 2020;Liang et al., 2020). The aim of the study is to extend these previous findings and develop a predictive risk score based on demographic, comorbidity, medication record and laboratory data using territory-wide electronic health records, without clinical parameters or imaging results. We hypothesized that incorporation of test results on successive time points would improve risk prediction.

## Methods

### Study design and population

This study was approved by the Institutional Review Board of the University of Hong Kong/Hospital Authority Hong Kong West Cluster. This was a retrospective, territory-wide cohort study of patients undergoing COVID-19 RT-PCR testing between 1^st^ January 2020 and 22^nd^ August 2020 in Hong Kong. The patients were identified from the Clinical Data Analysis and Reporting System (CDARS), a territory-wide database that centralizes patient information from individual local hospitals to establish comprehensive medical data, including clinical characteristics, disease diagnosis, laboratory results, and drug treatment details. The system has been previously used by both our team and other teams in Hong Kong ^7-9^. Patients demographics, prior comorbidities, hospitalization characteristics before admission due to COVID-19, medication prescriptions, laboratory examinations of complete blood counts, biochemical tests, diabetes mellitus tests, cardiac function tests, c-reactive protein, and blood gas tests were extracted. The list of ICD-9 codes for comorbidity identification, codes of intubation procedure, medication prescriptions are detailed in the **Supplementary Tables 1 to 3**.

**Table 1.**
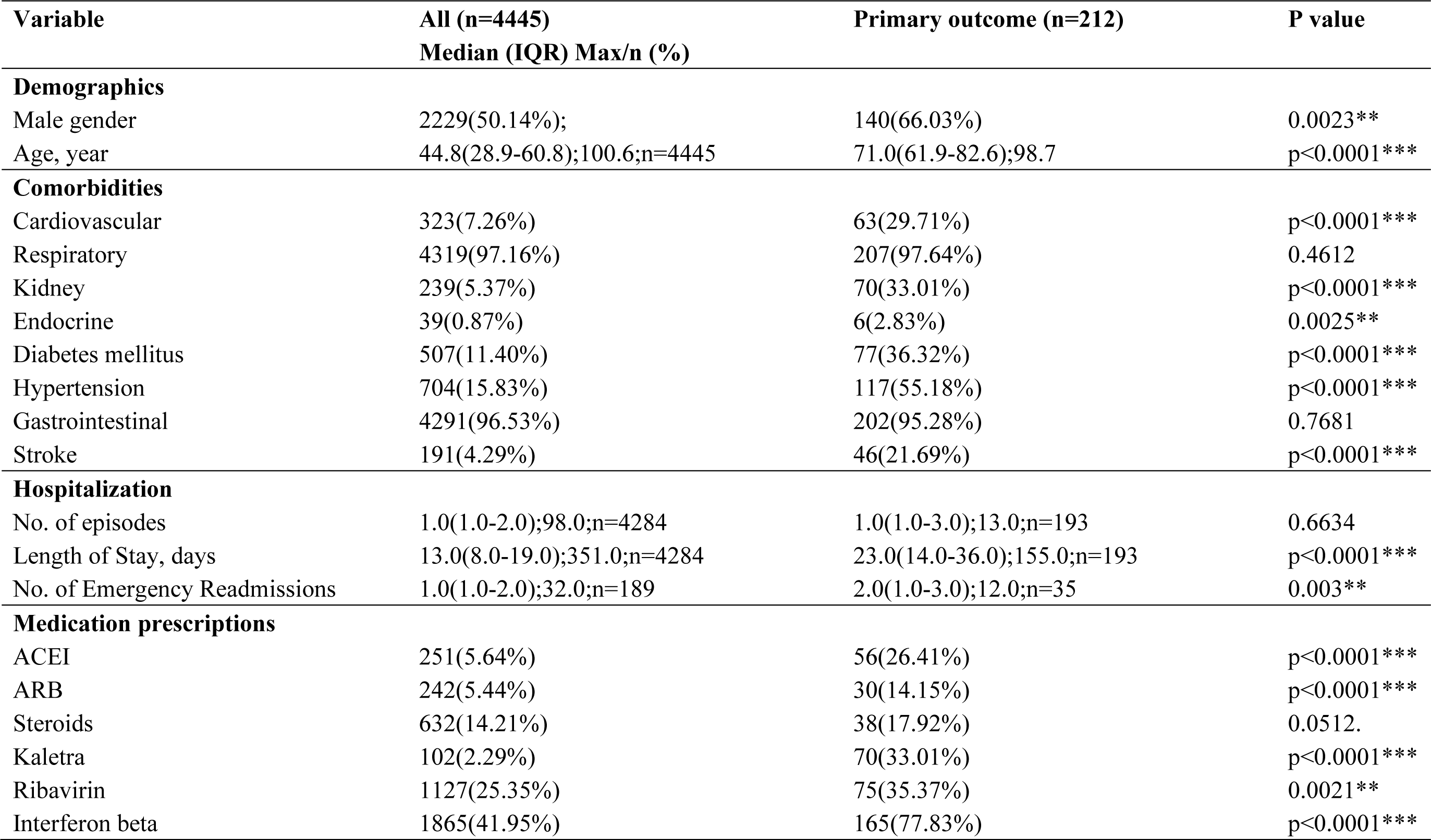
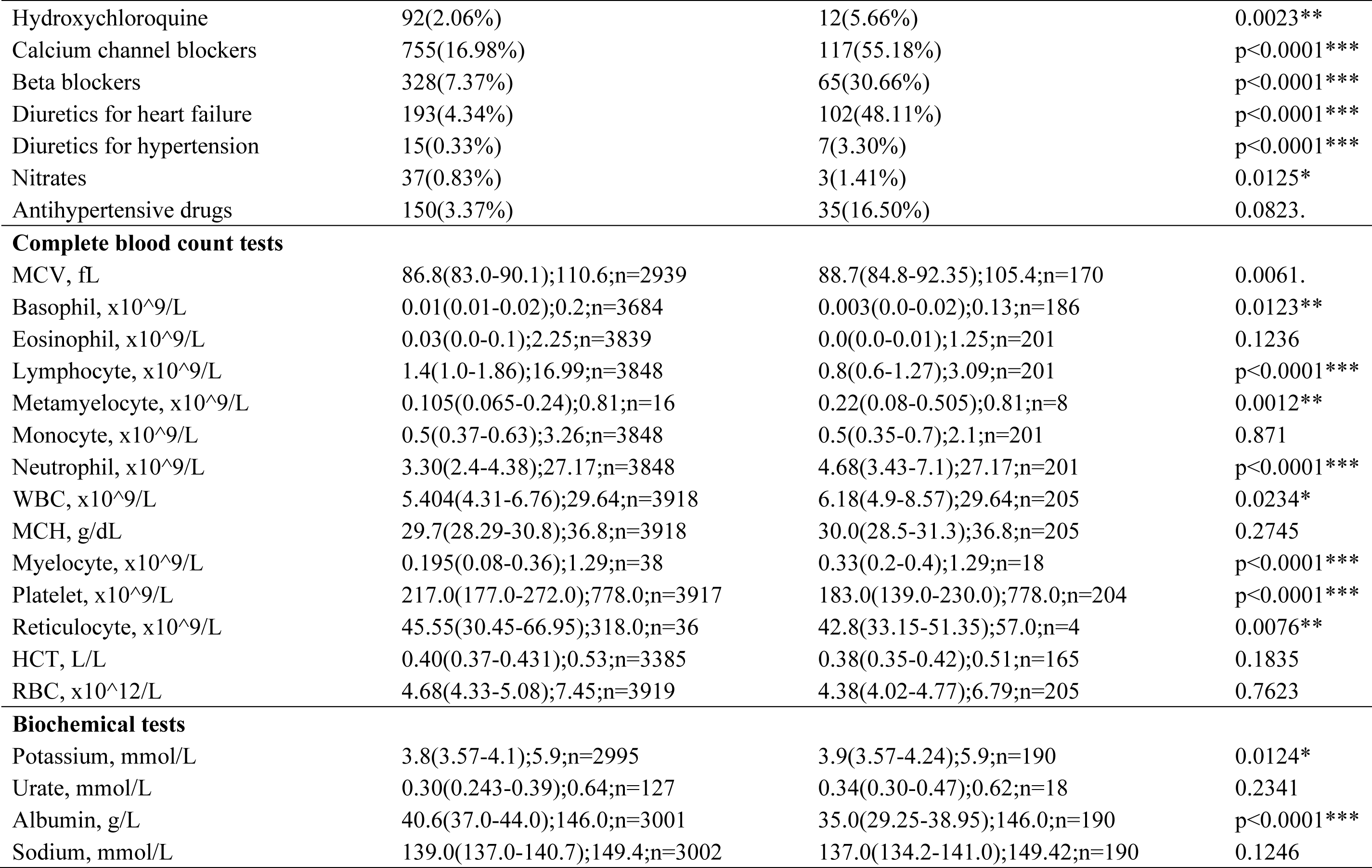
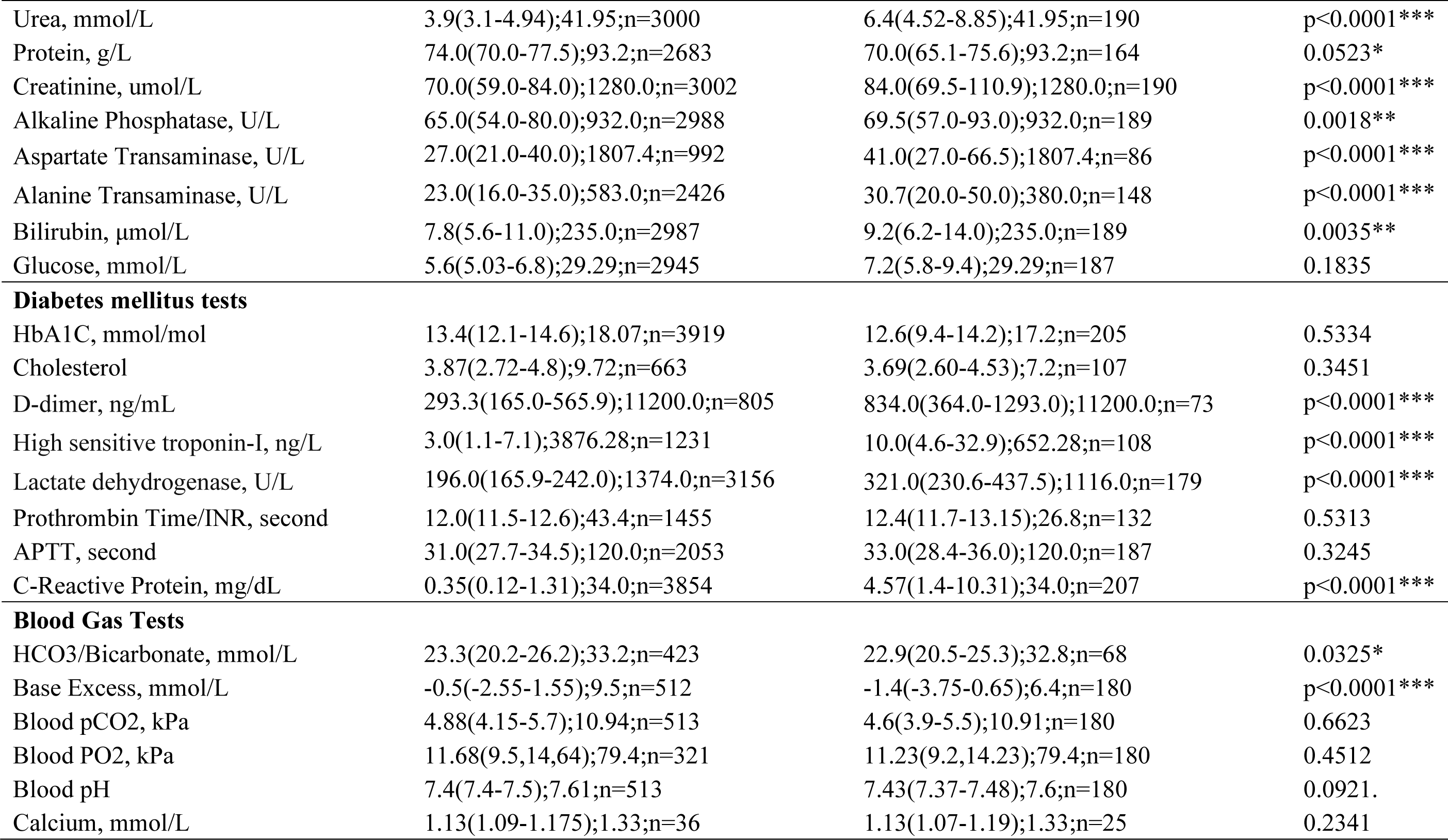
Baseline clinical characteristics of patients with COVID-19. Comparisons were made between patients meeting the primary outcome vs. those that did not. * for p≤ 0.05, ** for p ≤ 0.01, *** for p ≤ 0.001

### Outcomes and statistical analysis

The primary outcome was a composite of need for intensive care admission, intubation or all-cause mortality. Mortality data were obtained from the Hong Kong Death Registry, a population-based official government registry with the registered death records of all Hong Kong citizens linked to CDARS. The need for ICU admission and intubation were extracted directly from CDARS. Descriptive statistics are used to summarize baseline clinical characteristics of all patients with COVID-19 and based on the occurrence of the primary outcome. Continuous variables were presented as median (95% confidence interval [CI] or interquartile range [IQR]) and categorical variables were presented as count (%). The Mann-Whitney U test was used to compare continuous variables. The χ2 test with Yates’ correction was used for 2×2 contingency data. Univariate Cox regression identifies significant mortality risk predictors, which were used as input of multivariate Cox regression. Hazard ratios (HRs) with corresponding 95% CIs and p values were reported. An easy-for-use predictive model was developed using the beta coefficients of the multivariate Cox regression. Successive laboratory tests at least 24 hours apart were used. All statistical tests were two-tailed and considered significant if p value<0.001. They were performed using RStudio software (Version: 1.1.456) and Python (Version: 3.6).

## Results

### Basic characteristics

A total of 4445 patients (median age 44.8 years old, 95% CI: [28.9, 60.8]); 50% male) were diagnosed with the COVID-19 infection between 1^st^ January 2020 and 22^nd^ August 2020 in Hong Kong public hospitals or their associated ambulatory/outpatient facilities (**Table 1**). On follow-up until 8^th^ September 2020, a total of 212 patients (4.77%) met the primary outcome of need for intensive care admission or intubation, or death. The survival curve is presented in **Figure 1A**. Details on Cox regression analyses are shown in the **Supplementary Tables 4 and 5**.

**Figure 1A.**
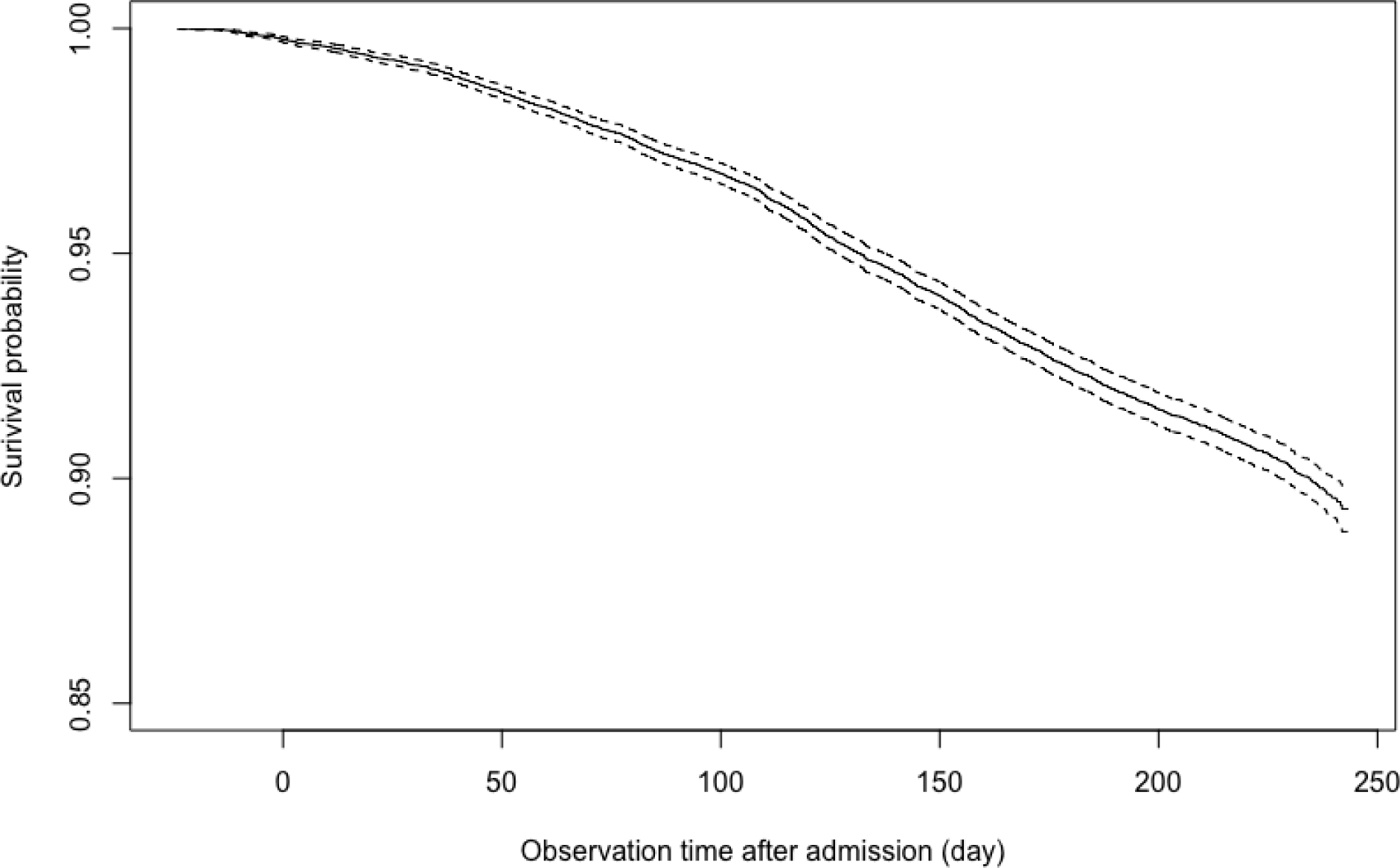
Survival curve of COVID-19 patient for the primary outcome, a composite of intensive care admission, need for intubation or death.

### Development of a clinical risk score and validation

For clinical practice, it is impractical to precisely input the values of all variables assessed from the different domains of the health records. We built a simple, easy-to-use model based on the number of abnormalities observed in each domain (**Table 2**). Using Harrell’s C-index and the area under the receiver operator characteristic curve (AUC) as performance evaluation metrics, we compared the prediction strengths of different criteria for the clinical risk score using a five-fold cross validation approach. The first model required cut-off values of different variables at baseline, and laboratory examinations on for each successive 24 hours was compared to cut-off to determine whether the criterion was met at each time point (**Supplementary Table 6**). In the second model, the criterion was met if the value was abnormal by standard laboratory criteria, without using optimal cut-off values (**Supplementary Table 7**). In the third model, laboratory test results are compared to the criteria without cut-off values, to see if they were met on successive testing (**Supplementary Table 8**). For example, if a particular criterion is met on day 1, then they will automatically fulfill the criteria for subsequent days.

**Table 2.** Easy-to-use score system for early prediction of severe COVID-19 disease.

| **Risk factor** | **Cut-off** | **Score** |
| --- | --- | --- |
| Male gender | Male | 4 |
| Age | 60-64 years old | 2 |
|  | 65-69 years old | 3 |
|  | 70-74 years old | 3 |
|  | ≥75 years old | 10 |
| Hypertension or on any anti-hypertensive drug | Present | 1 |
| Stroke | Present | 2 |
| Diabetes mellitus | Present | 1 |
| Myocardial infarction, heart failure or on any medication for these conditions | Present | 4 |
| Respiratory disease | Present | 3 |
| Kidney disease | Present | 2 |
| High neutrophil | 5.44 x10<sup>9</sup>/L | 1 |
| High monocyte | 0.80 x10<sup>9</sup>/L | 1 |
| Low lymphocyte | 0.77 x10<sup>9</sup>/L | 2 |
| High sodium | 136.4 mmol/L | 2 |
| High potassium | 4.32 mmol/L | 1 |
| High urea | 20.35 mmol/L | 5 |
| High ALT | 56 U/L | 1 |
| High ALP | 143 IU/L | 1 |
| High sensitive troponin-I, ng/L | 7.6 ng/L | 1 |
| High prothrombin time/INR | 12.6 seconds | 1 |
| High APTT | 38.8 seconds | 2 |
| High D-dimer | 831.46 ng/mL | 1 |
| High C-reactive protein | 0.88 mg/dL | 2 |
| Low base excess | -3.5 | 3 |
| Low bicarbonate | 28.3 | 2 |

Finally, we calculated the clinical risk score of each COVID-19 patient based on the third model and plotted the distribution of the whole cohort (**Figure 1B**). Patients meeting the primary outcome (n=212) have significantly higher risk score (median: 19, 95% CI: 14-24, max: 37) than those who did not (median: 8, 95% CI: 5-11, max: 34) **(Supplementary Table 9)**, indicating the performance of the clinical risk score (**Supplementary Table 10**). Receiver operating characteristic curve (ROC) of predicting adverse composite outcome of COVID-19 patients with the dichotomized risk score cut-off is shown in **Figure 1C**, demonstrating an AUC of 0.91. Further we generated the survival curves stratified by the dichotomized risk score in **Figure 1D**, where yellow and blue curves represent the survival analysis for patients with a clinical risk score is larger and smaller than the cut-off, respectively.

**Figure 1B.**
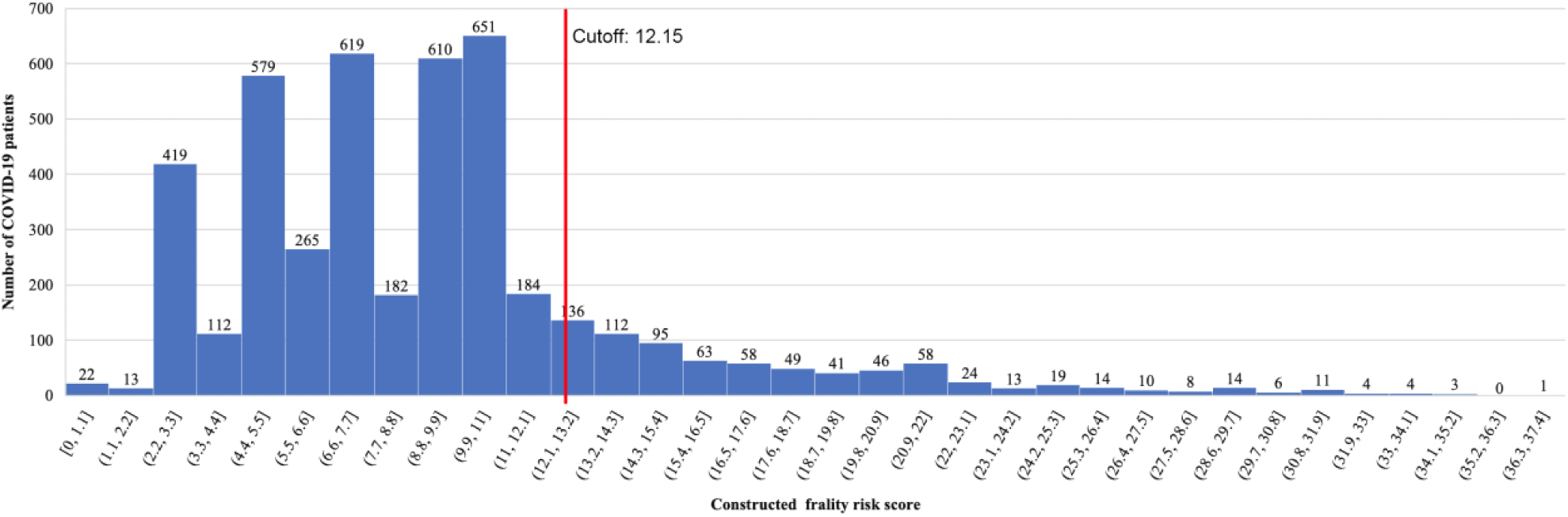
Distribution of derived risk score for composite outcome identification.

**Figure 1C.**
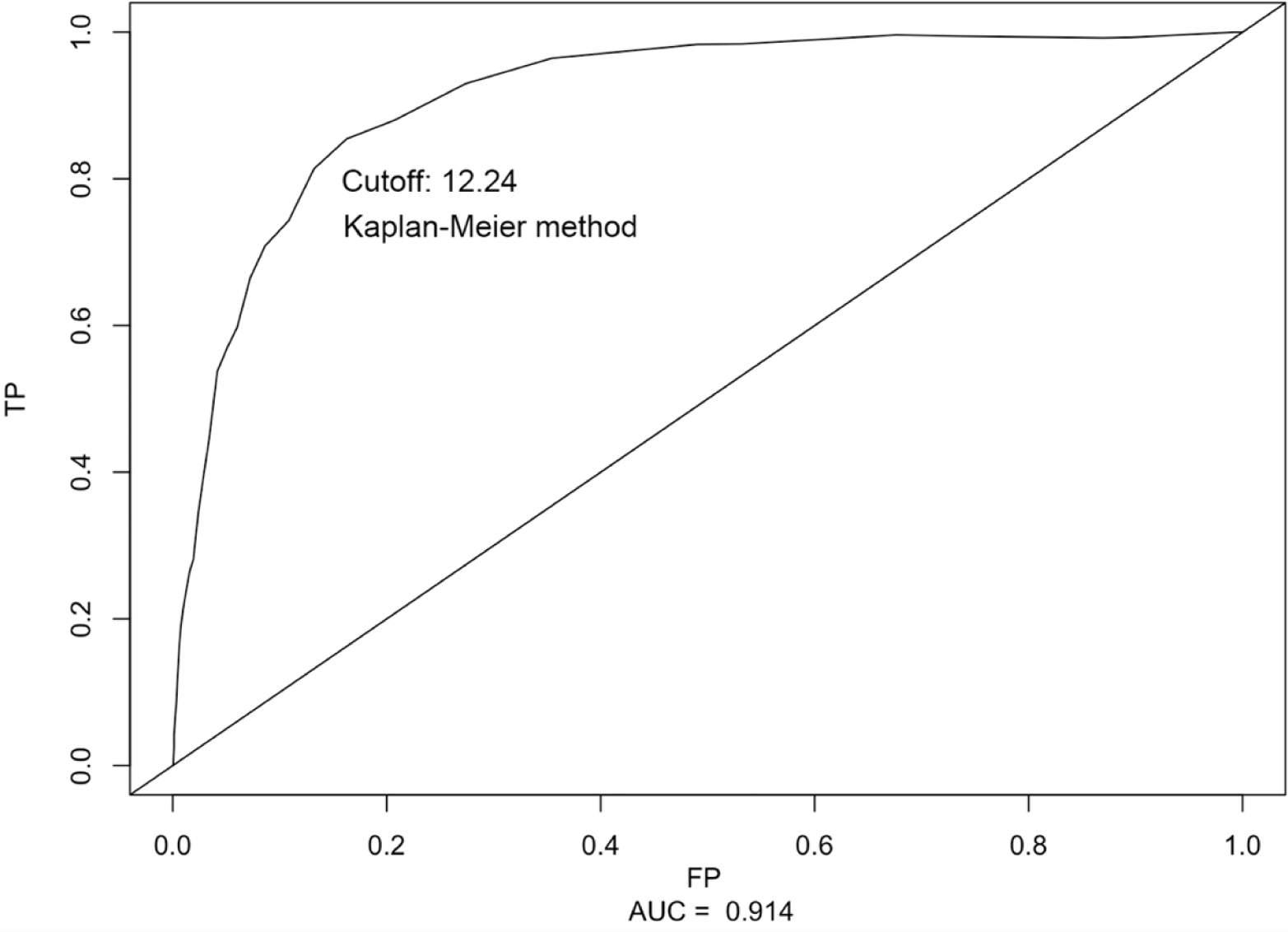
Receiver operating characteristic curve (ROC) of predicting adverse composite outcome of COVID-19 patients with dichotomized risk score.

**Figure 1D.**
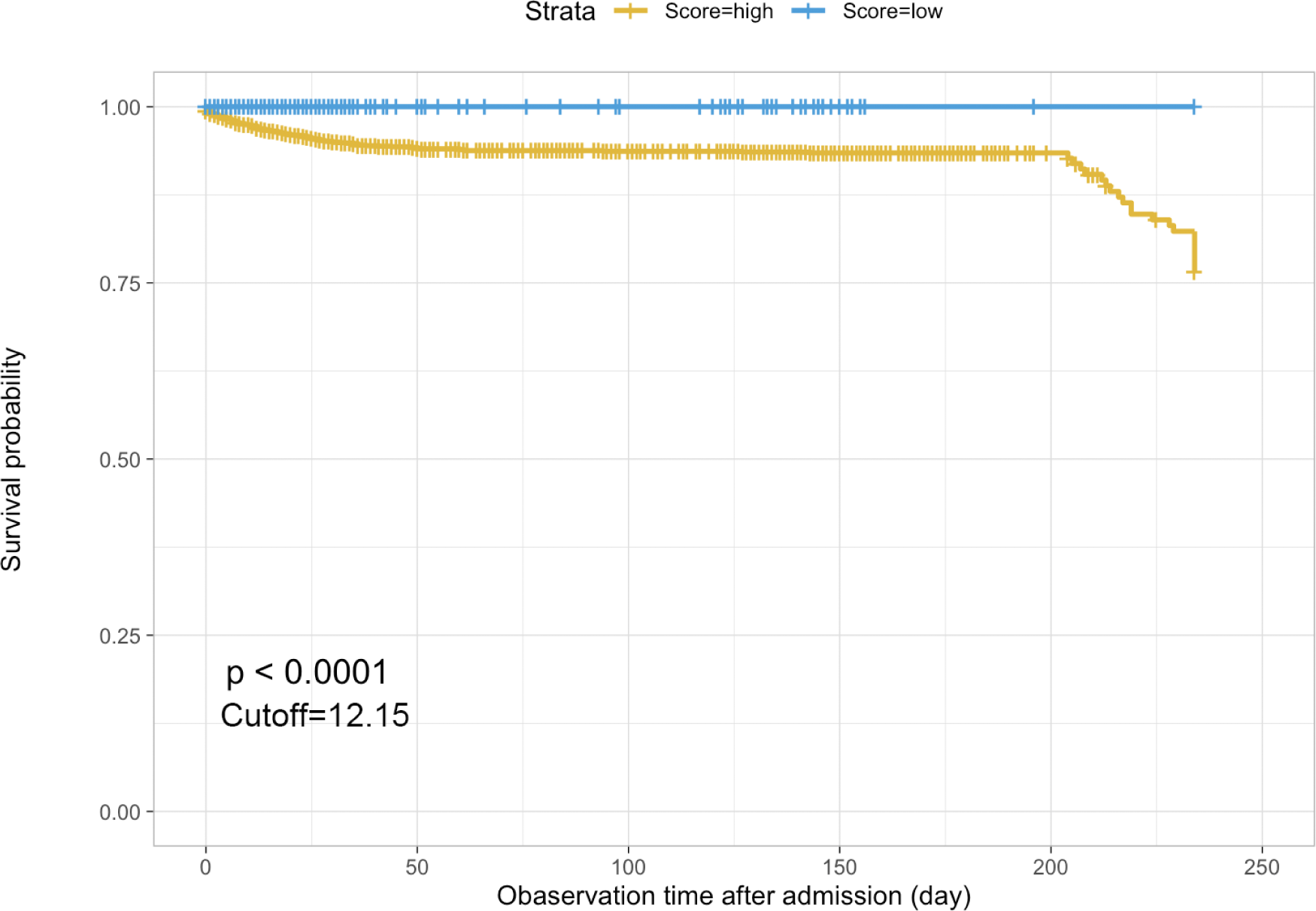
Survival curve of COVID-19 patients stratified by dichotomized risk score.

## Discussion

In this study, we developed a simple clinical score to predict severe COVID-19 disease based on age, gender, medical comorbidities, medication records, and laboratory examination results. This achieved good predictions with a c-statistic of 0.91, even without the consideration of clinical parameters such as symptoms, blood pressure, oxygen status on presentation or chest radiograph results.

COVID-19 disease has placed significant pressures on healthcare systems worldwide. Early risk stratification may better direct the use of limited resources and allow clinicians to triage patients and make clinical decisions based on limited evidence objectively. For example, low-risk patients may require simple monitoring only, whilst patients that are likely to deteriorate may benefit from intensive drug treatment or intensive care. Currently, the availability of simple clinical risk scores for risk stratification is limited. The COVID-GRAM predicts development of critical illness, based on symptoms, radiograph results, clinical and laboratory details (Liang et al., 2020). Similarly, the 4C Mortality Score included eight variables readily available at initial hospital assessment: age, sex, number of comorbidities, respiratory rate, peripheral oxygen saturation, level of consciousness, urea level, and C-reactive protein (score range 0-21 points) (Knight et al., 2020). These scores produced moderately accurate predictions with C-index values of 0.86 and 0.61-0.76, respectively. Our simple and easy-to-use model was based on comorbidity, drug, and laboratory data only, without needing clinical assessment details or the need of chest imaging. The model based on test results taken on the day of admission already demonstrated an excellent predictive value with a C-index of 0.87. Incorporation of test results on successive time points did not further improve risk prediction, indicating that initial data are sufficient to produce accurate predictions of severe disease.

### Limitations

The major limitation of this study is that it is based on a single territory-wide cohort. The model should be externally validated using patient data from other different regions.

## Conclusion

A simple clinical score based on only demographics, comorbidities, medication records and laboratory tests accurately predicted severe COVID-19 disease, even without including symptoms on presentation, blood pressure, oxygen status or chest radiograph results. The model based on test results taken on the day of admission showed an excellent predictive value. Incorporation of test results on successive time points did not further improve risk prediction.

## Data Availability

Data available upon request.

