## Supplementary Appendix for "Development of a predictive risk model for severe COVID-19 disease using population-based administrative data"

**Table 1. Drugs prescribed for COVID-19 patients**

| |
|---|
| **ACEI** |
| LISINOPRIL, PERINDOPRIL TERTBUTYLAMINE, PERINDOPRIL, RAMIPRIL, ENALAPRIL MALEATE, CAPTOPRIL, CAPTOPRIL TABLET, CAPTOPRIL, CAPTOPRIL TABLET 12.5MG--->, CAPTOPRIL, PERINDOPRIL ARGININE, PERINDOPRIL, CAPTOPRIL *FOR ORAL SOLUTION*, CAPTOPRIL, PERINDOPRIL, MALEATE, ARGININE |
| **ARB** |
| LOSARTAN POTASSIUM, IRBESARTAN, TELMISARTAN, CANDESARTAN CILEXETIL, CANDESARTAN, IRBESARTAN 300MG + HYDROCHLOROTHIAZ, IRBESARTAN, MICARDIS PLUS 40/12.5 (OR EQUIV), MICARDIS, MICARDIS PLUS 40/12.5 (OR EQUIV), MICARDIS, CO-DIOVAN 80/12.5 (OR EQUIV), DIOVAN, VALSARTAN, IRBESARTAN 150MG + HYDROCHLOROTHIAZ, IRBESARTAN, LOSARTAN K 50MG + HYDROCHLOROTHIAZI, LOSARTAN, LOSARTAN K 100MG + HYDROCHLOROTHIAZ, CO-DIOVAN 160/12.5 (OR EQUIV), SPARSENTAN/IRBESARTAN (CLINICAL TRI, IRBESARTAN, LOSARTAN OWN STOCK, LOSARTAN, CILEXETIL |
| **Steroid** |
| PREDNISOLONE, PROMETHAZINE COMPOUND, PROMETHAZINE HCL, PREDNISOLONE (SODIUM PHOSPHATE), METHYLPREDNISOLONE SODIUM SUCCINATE |
| **Kaletra** |
| KALETRA 100/25 (OR EQUIV), KALETRA (OR EQUIV) |
| **RIBAVIRIN** |
| **RIFAMPICIN** |
| **INTERFERON BETA-1B** |
| **HYDROXYCHLOROQUINE** |
| HYDROXYCHLOROQUINE SULPHATE |
| **Calcium channel blockers** |
| AMLODIPINE (BESYLATE), OLMESARTAN/NORVASC 40MG/5MG, DILTIAZEM HCL, NIFEDIPINE, VERAPAMIL HCL, FELODIPINE |
| **Beta blockers** |
| ATENOLOL, BISOPROLOL FUMARATE, METOPROLOL TARTRATE, METOPROLOL, CARVEDILOL, PROPRANOLOL HCL |
| **Diuretics for heart failure** |
| SPIRONOLACTONE, FRUSEMIDE (FUROSEMIDE), METOLAZONE |

**Diuretics for hypertension**
AMILORIDE HCL, HYDROCHLOROTHIAZIDE, INDAPAMIDE (NATRILIX SR) SUSTAINED, INDAPAMIDE, DYAZIDE (OR EQUIV)
**Nitrates**
ISOSORBIDE DINITRATE, ISOSORBIDE, DINITRATE
**Other Antihypertensive drugs**
METHYLDOPA, DOXAZOSIN (MESYLATE) GITS, TERAZOSIN HCL

**Table 2. Codes for Comorbidities of COVID-19 Patients**

| Comorbidity | Codes |
|---|---|
| Respiratory | 786.09, 518.81, 780.53, 137, E912, 465.9, 518.81, 518.81, 79.6, 518.81, 519.8, 780.59, 799.1, 780.57, 518.82, 480.1, 786.3, 519.8, 997.3, 165.9, 519.9, 648.91, 162.9, 162.3, 197, 162.5, 162.4, 486, 518.89, 496, 162.8, 415.1, V10.11, 518, 162.9, 11.96, 482.1, 507, 513, 11.9, 511.8, 511.1, 11.94, 516.8, 793.1, 482.4, 507, 515, 197, 11.93, 482, 482.83, 518.4, 482.3, 482.2, 415.1, 502, 518.89, 235.7, 793.1, 934.8, 516.9, 136.3, 38.49, 506, 112.4, 487, 481, 117.9, 38.2, 518, 11.95, 79.89, 518.1, 480.9, 505, 516.8, 495.9, 518.3, 11.23, 416.8, 513, 397.1, 117.3, 483, 508, 998.81, 416, 514, 861.21, 502, 934.8, 480.8, 648.93, 11.2, 492.8, 484.6, 78.5, 484.1, 516.3, 415.1, 416.9, 415, 416.9, 429.89, 415, 747.49, 745, 417, 770.7, 427.5, 416.9, 416, 416.8, 746.02, 573.8, 642.9, 416, 747.3, 747.3, 770.3, 779.8, 515, 424.3, 416, 417.8, 747.3, 747.3, 745.4, 518.81, 786.09, V12.6, 478, 748.5, 162.9, 996.84, 748.5, 748.6, V42.1, 748.5, 11.05, 162, 518, 747.42, 518.89, 748.5, 517.2 |
| Kidney | 198.7, 189, 189, 585.9, V56.0, 189.1, 584.9, 189.1, 593.9, 189, 189, 189.8, 239.5, 189, 583.81, 593.9, 591, V10.52, 250.4, 591, 255.4, 590.8, 586, 585.1, 591, 189, 591, 239.7, 788, 996.39, 189.1, 588.9, 592, 996.39, 250.4, 593.2, 255.4, 572.4, 194, 255, 453.3, 198, 585.9, 996.39, 996.39, 590.1, 591, 591, 223, 591, 584.9, 585.9, 250.41, 996.39, 581.9, 227, 593.5, 583.89, 593.89, 404.93, 255.5, 788.9, 250.43, 227, 405.92, 592, 753.12, 996.39, E879.1, V42.0, 592, 580.89, 403.9, 593.9, V59.4, 585.9, 580.9, 250.41, 996.39, 585.9, 794.4, 584.8, 584.5, 255.9, 441.4, V58.49, 404.11, 593.9, 592, 585, 759.1, 753.11, E879.1, 753.15, 585.9, 274, 588.8, 403.91, 404.9, 404.91, 404.92, 403, 404.01, 779.8, 250.4, 227, 227, 227, 404.93, 996.81, 593.89, 753.8, 593.2, 592, 753, 996.81, 223, 589.1, 996.81, 582.9, 585.9, 996.81, 753.1, 753.3, E878.0, 593.9, 753.3, 753.17, 583.9, 593.9, 589, 866 |
| Endocrine | 202.8, 200.1, 200.12, 201.9, 204, 202.88, 200.18, 196, 204.01, 785.6, 200.11, 200.13, 202.8, 785.6, 202.85, 202.81, 202.8, 202.8, 196, 196.9, 202.8, 202.82, 785.6, 202.8, 202, 202.87, V10.79, 785.6, 202.84, 202.01, 196.8, 457.1, 12.1, 785.6, V10.79, 196.5, 196.2, 238.7, 196.1, 200.14, 457.2, 238.7, V10.61, 201.9, 457, 202.8, 289.3, 245.2, 238.7, 785.6, 457.9, 785.6, 202.93, 196.9, 202.97, 757, V10.71, 288.8, 204.1, 202.83, 457.1, 289.3, 785.6, V77.9, 237.4, 239.7, 198.89, 623.5, 259.9, 200.2, V10.71 |
| Diabetes mellitus | 251.2, 362.01, 362.02, 250.4, 250.82, 790.2, 790.6, 250.5, 250.5, 250.6, 357.2, 790.2, 250, 250.4, 250.6, 250.8, 250.51, 250.5, 250.8, 250.82, 250.51, 250.51, V77.1, 250.12, 251.2, 250.12, 250.5, 250.83, 251.2, 250.41, 251.1, 250.52, 250.5, 648.81, 250.43, 250.53, 250.53, 250.81, 250.22, 250.13, 250.22, 250.83, 250.41, 250.5, 250.52, 250.52, 250.82, 253.5, V18.0, 588.1 |

| | |
|---|---|
| Hypertension | 401.9, 401.9, 250.82, 790.6, 401.9, 401.9, 250.82, 796.2, 402.9, 250.83, 405.99, 642.93, 642.01, 642.91, 401.9, E942.6, 405.09, 403.9, 437.2, 401, 401.1, 401, 642.33, 348.2, 779.8, 365.04, 572.3, 416, 416, 405.91, 416.8, 642.3 |
| Gastrointestinal | 153.3, 154.1, 153.9, 569.89, 154, 153.1, 578.9, 560.9, 569.3, 537.89, 558.9, 562.1, 153.6, 239, 532.3, 569.89, 532.7, 535.6, 558.9, 38.42, 569.89, 8.45, 153.2, 569.49, 79.89, 532.9, V58.11, 569, 154.1, 41.4, 537.89, 152.1, 578.9, V10.05, 787.8, 197.4, 535.5, V10.06, 9, 569.83, 569.6, 153.4, 560.9, 537.3, 41.04, 569.84, 239, 569.81, 8.8, 535, 560.9, 532, V45.89, V12.72, 532.4, V10.09, 560.81, 235.2, 38.49, 8.45, 235.2, 532.9, 569.81, 537.89, 557.9, 569.41, 997.4, 14.8, 787.99, 8.46, 535.5, 569.41, 997.4, 578.9, 569.82, 537.9, 560.1, 569.82, 557.9, 211.3, 556.9, 562, 558.9, 578.9, 536.9, 8.46, 535.6, 566, V71.9, 569.49, 564.3, V44.4, 569.89, 564.8, 8.46, 569.83, 997.4, 997.4, 997.4, 562.11, 211.2, 9.1, 211.3, 8.47, 8.5, 211.3, 569.83, 532.1, 535.61, 560, 569.83, 565.1, 619.1, 152.9, 568, 566, 569.43, 152, 8.46, 562.11, 8.61, 569.83, 569.83, 569.81, 596.1, 535.5, 151.4, 151.9, 151.5, 151.8, 151.1, 456.8, 531.7, 535.4, 531.3, V15.2, 537.89, 211.1, 531.9, V10.04, 235.2, 531, 531.4, 456.8, 535.1, 151.3, 230.2, 151.6, 535, 211.1, 536.3, 535, V10.04, 535.51, 578.9, 531.1, 535.1, 456.8, 531.5, 537.84, 535.01, 530.7, 535.1, 535.2, 535.5, 537.6, 202.83, 535.1, 531.4, 558.9, 558.9, 558, 569.85, 153, 555.1, 562.1, 562.13, 562.11, 562.12, 569.83, V76.49, 560.2, 230.4, 569.3, 154 |
| Stroke | 434.11, 434.91, 433.01, 433.11, 433.21, 433.31, 433.81, 433.91, 434.01, 434.91, 436, 435, 430, 431, 432, 432.1, 432.9, 853, 852, 852.01, 852.02, 852.03, 852.04, 852.05, 852.06, 852.07, 852.08, 852.09, 852.2, 852.21, 852.22, 852.23, 852.24, 852.25, 852.26, 852.27, 852.28, 852.29, 852.4, 852.41, 852.42, 852.43, 852.44, 852.45, 852.46, 852.47, 852.48, 852.49, I64, 992, 992, 434.01, 433, 331, 851, 434, 434.1, 348.5, 747.81, 414.9, 457.9, 720, 681.1, 473.9, 290.3, 524.02, 720, 414.9, 215.3 |

**Table 3. Codes for Identifying Patients Needing Intubation**

| **Procedure** | **Codes and Description** |
|---|---|
| Intubation | Cont invasive mech vent->96 hours (96.72:0) |
| | Cont invasive mech vent-<96 hours (96.71:0) |
| | Invasive mechanical ventilation (96.70:0) |
| | Endotracheal intubation (96.04:0) |
| | Respiratory tract intubation (96.05:0) |

**Supplementary Table 4. Univariate Cox regression to identify significant risk predictors of the primary outcome (composite outcome of intensive care admission, intubation or death).**

* for p≤ 0.05, ** for p ≤ 0.01, *** for p ≤ 0.001

| **Variable** | **Composite outcome HR [95% CI]** | **P value** |
|---|---|---|
| Male gender | 1.9 [1.4, 2.6] | <0.0001*** |
| Age, year | | |
| 60-64 | 1.45 [1.22, 2.32] | 0.0015** |
| 65-69 | 2.12 [1.76, 3.31] | <0.0001*** |
| 70-74 | 3.19 [2.45, 5.13] | <0.0001*** |
| >=75 | 12.41 [8.09, 16.34] | <0.0001*** |
| Cardiovascular | 5.5 [4.1, 7.4] | <0.0001*** |
| Respiratory | 5.5 [2.1, 14.1] | 0.0004*** |
| Kidney | 9.1 [6.8, 12.1] | <0.0001*** |
| Endocrine | 4.1 [1.8, 9.1] | 0.0007*** |
| Diabetes mellitus | 4.5 [3.4, 6.0] | <0.0001*** |
| Hypertension | 6.9 [5.2, 9.0] | <0.0001*** |
| Gastrointestinal | 2.7 [1.3, 5.4] | 0.0067** |
| Stroke | 6.1 [4.4, 8.4] | <0.0001*** |
| No. of hospitalizations | 1.0 [1.0, 1.1] | <0.0001*** |
| Length of Stay, day | 1.0 [1.0, 1.0] | <0.0001*** |
| No. of emergency readmissions | 1.1 [1.0, 1.1] | 0.0578. |
| ACEI | 5.9 [4.3, 8.0] | <0.0001*** |
| ARB | 2.6 [1.8, 3.9] | <0.0001*** |
| Steroids | 1.4 [1.0, 2.0] | 0.0641. |
| Kaletra | 34.0 [25.3, 45.7] | <0.0001*** |
| Ribavirin | 1.7 [1.3, 2.2] | 0.0005*** |
| Interferon beta | 9.1 [6.0, 13.7] | <0.0001*** |
| Hydroxychloroquine | 2.6 [1.5, 4.7] | 0.0013** |
| Calcium channel blockers | 5.9 [4.5, 7.7] | <0.0001*** |
| Beta blockers | 5.5 [4.1, 7.4] | <0.0001*** |
| Diuretics for heart failure | 20.0 [15.2, 26.4] | <0.0001*** |
| Diuretics for hypertension | 10.0 [4.7, 21.4] | <0.0001*** |
| Nitrates | 1.5 [0.5, 4.6] | 0.515 |
| Antihypertensive drugs | 5.217[3.62, 7.52] | <0.0001*** |
| MCV, fL | 1.046 [1.022, 1.070] | 0.0001*** |
| Basophil, x10^9/L | 0.048 [0.03, 43.7] | 0.383 |
| Eosinophil, x10^9/L | 0.23 [0.14, 0. 28] | <0.0001*** |
| Lymphocyte, x10^9/L | 0.2 [0.1, 0.3] | <0.0001*** |
| Metamyelocyte, x10^9/L | 2.0 [0.1, 30.7] | 0.606 |
| Monocyte, x10^9/L | 2.1 [1.3, 3.4] | 0.004** |
| Neutrophil, x10^9/L | 1.3 [1.2, 1.3] | <0.0001*** |

| | | |
|---|---|---|
| WBC, x10^9/L | 1.2 [1.2, 1.2] | <0.0001*** |
| MCH, g/dL | 1.1 [1.0, 1.1] | 0.0185* |
| Myelocyte, x10^9/L | 18.0 [2.8, 114.1] | 0.0022** |
| Platelet, x10^9/L | 1.0 [1.0, 1.0] | <0.0001*** |
| Reticulocyte, x10^9/L | 1.0 [1.0, 1.0] | 0.673 |
| HCT, L/L | 0.56 [0.34, 0.87] | <0.0001*** |
| RBC, x10^12/L | 0.4 [0.3, 0.5] | <0.0001*** |
| K/Potassium, mmol/L | 1.7 [1.2, 2.4] | 0.0015** |
| Urate, mmol/L | 8.8 [0.2, 377.3] | 0.258 |
| Albumin, g/L | 1.0 [1.0, 1.0] | 0.15 |
| Na/Sodium, mmol/L | 0.9 [0.9, 0.9] | <0.0001*** |
| Urea, mmol/L | 1.1 [1.1, 1.1] | <0.0001*** |
| Protein, g/L | 0.9 [0.9, 0.9] | <0.0001*** |
| Creatinine, umol/L | 1.004 [1.003, 1.005] | <0.0001*** |
| Alkaline Phosphatase, U/L | 1.003 [1.001, 1.005] | 0.0004*** |
| Aspartate Transaminase, U/L | 1.002 [1.001, 1.003] | <0.0001*** |
| Alanine Transaminase, U/L | 1.007 [1.004, 1.009] | <0.0001*** |
| Bilirubin, µmol/L | 1.025 [1.019, 1.031] | <0.0001*** |
| Glucose, mmol/L | 1.2 [1.1, 1.2] | <0.0001*** |
| HbA1c, mmol/mol | 0.935 [0.912, 0.958] | <0.0001*** |
| Cholesterol, mmol/L | 0.877 [0.779, 0.988] | 0.0311* |
| D-dimer, ng/mL | 1.000 [1.000, 1.000] | <0.0001*** |
| High sensitive troponin-I, ng/L | 1.001 [1.000, 1.001] | 0.0388* |
| Lactate dehydrogenase, U/L | 1.004 [1.004, 1.005] | <0.0001*** |
| Prothrombin Time/INR, second | 1.1 [1.0, 1.1] | <0.0001*** |
| APTT, second | 1.0 [1.0, 1.1] | <0.0001*** |
| C-Reactive Protein, mg/dL | 1.159 [1.142, 1.176] | <0.0001*** |
| **Blood Gas Tests** | | |
| HCO3/Bicarbonate, mmol/L | 0.987 [0.92, 0.999] | <0.0001*** |
| Base Excess, mmol/L | 0.911 [0.878, 0.945] | <0.0001*** |
| Blood pCO2, kPa | 0.902 [0.797, 1.020] | 0.1023 |
| Blood PO2, kPa | 0.65 [0.21, 1.031] | 0.2314 |
| Blood pH | 0.185 [0.028, 1.210] | 0.0782. |
| Calcium, mmol/L | 0.195 [0.003, 13.4] | 0.449 |

**Supplementary Table 5. Multivariate analysis of significant risk predictors of the composite outcome.**

* for p≤ 0.05, ** for p ≤ 0.01, *** for p ≤ 0.001

| Variable | Composite outcome HR [95% CI] | P value |
|---|---|---|
| Male gender | 3.57 [1.14, 10.53] | 0.0012** |
| Age, year | | |
| 60-64 | 1.57 [1.01, 2.32] | 0.0253* |
| 65-69 | 2.46 [1.67, 3.63] | <0.0001*** |
| 70-74 | 2.59 [1.70, 3.94] | <0.0001*** |
| >=75 | 10.17 [7.72, 13.42] | <0.0001*** |
| Cardiovascular | 3.2 [2.1, 5.2] | <0.0001*** |
| Respiratory | 2.8 [1.2, 3.6] | <0.0001*** |
| Kidney | 2.4 [1.3, 3.2] | <0.0001*** |
| Diabetes mellitus | 1.4 [1.2, 2.7] | <0.0001*** |
| Hypertension | 1.4 [1.2, 1.2] | <0.0001*** |
| Stroke | 1.4 [1.2, 2.2] | <0.0001*** |
| No. of hospitalizations | 1.4 [1.2, 2.5] | <0.0001*** |
| Length of Stay | 0.8 [0.6, 0.8] | <0.0001*** |
| No. of emergency readmissions | 1.4 [1.1, 2.2] | <0.0001*** |
| ACEI | 1.6 [1.2, 2.1] | <0.0001*** |
| ARB | 1.8 [1.3, 2.5] | <0.0001*** |
| Kaletra | 1.1 [1.0, 1.6] | <0.0001*** |
| Ribavirin | 1.2 [1.1, 1.4] | <0.0001*** |
| Interferon beta | 1.5 [1.3, 1.6] | <0.0001*** |
| Calcium channel blockers | 1.3 [1.1, 2.2] | 0.0351* |
| Beta blockers | 1.6 [1.1, 2.7] | 0.0286* |
| Diuretics for heart failure | 1.7 [1.1, 2.5] | <0.0001*** |
| Diuretics for hypertension | 2.5 [1.2, 2.6] | <0.0001*** |
| Antihypertensive drugs | 1.4 [1.1, 1.9] | <0.0001*** |
| MCV, fL | 1.2 [1.1, 1.5] | <0.0001*** |
| Eosinophil, x10^9/L | 0.8 [0.5, 0.9] | 0.4561 |
| Lymphocyte, x10^9/L | 0.3 [0.2, 0.7] | 0.0036** |
| Monocyte, x10^9/L | 1.4 [1.1, 2.2] | <0.0001*** |
| Neutrophil, x10^9/L | 1.4 [1.2, 2.6] | <0.0001*** |
| WBC, x10^9/L | 2.2 [1.5, 12.5] | 0.2314 |
| Platelet, x10^9/L | 1.2 [1.1, 2.09] | <0.0001*** |
| HCT, L/L | 0.7 [0.3, 0.9] | <0.0001*** |
| RBC, x10^12/L | 1.3 [1.0, 2.1] | <0.0001*** |
| Potassium, mmol/L | 1.2 [1.1, 2.1] | <0.0001*** |
| Sodium, mmol/L | 1.6 [1.1, 2.6] | <0.0001*** |
| Urea, mmol/L | 5.2 [1.5, 18.5] | <0.0001*** |
| Protein, g/L | 1.5 [1.2, 2.9] | <0.0001*** |
| Creatinine, umol/L | 1.3 [1.1, 2.2] | <0.0001*** |
| Alkaline Phosphatase, U/L | 1.3 [1.1, 2.1] | <0.0001*** |
| Glucose, mmol/L | 0.8 [0.6, 1.0] | <0.0001*** |
| HbA1c | 1.4 [1.2, 1.8] | <0.0001*** |
| Cholesterol, mmol/L | 1.6 [1.2, 2.5] | <0.0001*** |
| D-dimer, ng/mL | 1.1 [1.0, 1.5] | <0.0001*** |

| | | |
|---|---|---|
| High sensitive troponin-I, ng/L | 1.3 [1.2, 1.9]] | <0.0001*** |
| Lactate dehydrogenase, U/L | 1.3 [1.1, 1.8] | 0.0023** |
| Prothrombin Time/INR, second | 1.7 [1.2, 2.5] | <0.0001*** |
| APTT, second | 1.5 [1.3, 2.6] | 0.0006*** |
| C Reactive Protein | 1.7 [1.3, 2.6] | <0.0001*** |
| Base Excess | 2.7 [1.5, 3.6] | <0.0001*** |
| HCO3/Bicarbonate | 1.5 [1.2, 2.8] | <0.0001*** |

**Supplementary Table 6.** Prediction strength of laboratory tests on successive days using baseline cut-off values

| | Baseline (1st test) | 2nd test | 3rd test | 4th test | 5th test | 6th test | 7th test | 8th test |
|---|---|---|---|---|---|---|---|---|
| AUC | 0.91 | 0.90 | 0.85 | 0.87 | 0.87 | 0.86 | 0.82 | 0.83 |
| C-index | 0.87 | 0.86 | 0.86 | 0.85 | 0.86 | 0.82 | 0.78 | 0.77 |

**Supplementary Table 7.** Prediction strength of laboratory tests on successive days without using cut-off values

| | Baseline (1st test) | 2nd test | 3rd test | 4th test | 5th test | 6th test | 7th test | 8th test |
|---|---|---|---|---|---|---|---|---|
| AUC | 0.89 | 0.89 | 0.89 | 0.89 | 0.89 | 0.90 | 0.90 | 0.90 |
| C-index | 0.86 | 0.86 | 0.86 | 0.87 | 0.87 | 0.87 | 0.88 | 0.88 |

**Supplementary Table 8.** Prediction strength of laboratory tests cumulatively without using cut-off values

| | Baseline (1st test) | 2nd test | 3rd test | 4th test | 5th test | 6th test | 7th test | 8th test |
|---|---|---|---|---|---|---|---|---|
| AUC | 0.88 | 0.90 | 0.90 | 0.90 | 0.91 | 0.91 | 0.91 | 0.91 |
| C-index | 0.87 | 0.87 | 0.87 | 0.88 | 0.88 | 0.88 | 0.88 | 0.88 |

**Supplementary Table 9.** Derived score characteristics of patients with/without composite outcome

* for p≤ 0.05, ** for p ≤ 0.01, *** for p ≤ 0.001

| | **No Composite (n=4233) Median (IQR); Max** | **Composite (n=212) Median (IQR); Max** | **P value** |
|---|---|---|---|
| Derived risk score | 8(6-11);37 | 24.71(17.11-35.67);37 | <0.0001*** |

**Supplementary Table 10.** Stratification performance of score and dichotomized score system

| | **Cut-off** | **HR (95% CI)** | **Z value** | **P-value** |
|---|---|---|---|---|
| Score | 12.15 | 1.21 (1.19-1.22) | 25.50 | <0.0001*** |
| Score≥12.15 | - | 24.71 (17.11-35.67) | 17.11 | <0.0001*** |
